# Family functioning and psychiatric outcomes in children and young people with intellectual and developmental disabilities caused by rare genetic mutations

**DOI:** 10.64898/2026.05.10.26352561

**Authors:** Josephine E Haddon, Jessica H Hall, IMAGINE ID, Jeremy Hall, Michael J Owen, Marianne B M van den Bree

**Author notes:** The members of IMAGINE-ID are listed in the acknowledgements. **Corresponding author**: Prof. Marianne van den Bree, Division of Psychological Medicine and Clinical Neurosciences, Centre for Neuropsychiatric Genetics and Genomics, Cardiff University, Cardiff, UK.

## Abstract

**Background:** A range of rare chromosomal micro-deletions or -duplications (Copy Number Variants - CNVs) are associated with high risk of neurodevelopmental and mental health conditions (ND-CNVs). There is great individual variability in outcomes, but we lack insights into the contributing social factors, including family functioning.

**Methods:** Caregivers of 598 children and young people (CYP) with a range of 16 ND-CNVs and 222 siblings without ND-CNVs (controls) completed questionnaires on overall family climate (cohesion and conflict) as well as caregiver-CYP relationship warmth and hostility and took part in a research diagnostic interview about CYPs’ psychiatric symptoms. CYPs’ intelligence quotient (IQ) was also measured.

**Results:** Comparisons with published data from neurotypical families indicated that families affected by ND-CNVs are characterised by higher family cohesion and conflict as well as lower caregiver-CYP warmth and hostility. Symptoms of oppositional defiant disorder reduced more steeply in CYP with ND-CNVs compared to controls with increasing family cohesion (interaction effect: β = -0.14, p = 4.65 × 10⁻²). In contrast, they rose more steeply with increasing family conflict (interaction effect: β = 0.18, p = 1.05 × 10⁻²). Furthermore, symptoms of mood disorder increased more steeply with increased caregiver-CYP hostility in CYP with ND-CNVs (interaction effect: β = 0.15, p = 4.55 × 10⁻²).

**Conclusions:** Raising a CYP with a rare genetic condition is challenging. Timely access to interventions that support caregivers in fostering a positive family environment may reduce behavioural difficulties in CYP, with subsequent benefits for family functioning.

## Introduction

### Neurodevelopmental risk copy number variants (ND-CNVs)

A range of rare copy number variants (CNVs); mutations involving micro-deletion or - duplication of genetic material, have been found to be strongly linked to neurodevelopmental conditions such as intellectual disability, autism and ADHD (referred to as neurodevelopmental CNVs or ND-CNVs) as well as other psychiatric outcomes such as anxiety and oppositional defiant disorder (ODD) (e.g., Chawner et al. 2021). There is, however, considerable variability in phenotypic outcome even in individuals with the same variant (Chawner et al. 2019). Understanding potentially modifiable factors that influence phenotypic variability in CYP with ND-CNVs is important for counselling and supporting families and provides insight into targets for early intervention.

### Family Environment

The family environment is crucial in shaping children’s neurodevelopment, including their cognitive skills and mental health (Bush et al. 2020). One important aspect of the family environment is the overall emotional atmosphere, characterized by the quality of interactions, emotional expressions, and relationships among family members (referred to as family functioning from here onwards). Family functioning can be considered both at the systemic level of the family unit as a whole (overall family climate, including cohesion and conflict - e.g., Moos and Moos 2009), and/or the quality of individual dyadic relationships within the family (i.e., warmth, hostility and control in caregiver-child relationships, e.g., Conger 1989).

### Family functioning, psychopathology and cognition

Associations have been reported between family functioning and psychiatric outcomes including internalising (anxiety, depression) and externalising behaviours (aggression, oppositional defiance) as well as cognitive function in CYP in both population-based (Chiang and Bai 2022; McKinney and Renk 2011; Rabinowitz et al. 2016) as well as clinical cohorts (Allen et al. 2014; Shashi et al. 2010).

Higher levels of family cohesion are associated with lower levels of externalising and internalizing symptoms (van Eickels et al. 2022) and improved adaptive anger regulation in CYP (Houltberg et al. 2012). Conversely, increased family conflict correlates with higher rates of internalizing (e.g., Rabinowitz et al. 2016) and externalizing (e.g., McKinney and Renk 2011), as well as neurodevelopmental symptoms (e.g., of autism, Chan and Leung 2020). Furthermore, caregiver-CYP hostility has been associated with increased internalizing and externalizing behaviours (Bayer et al. 2012), and poorer cognitive outcomes in CYP (Lam et al. 2018). Conversely, higher levels of caregiver-CYP warmth have been linked to increased emotional regulation skills (i.e. managing frustration, Isdahl-Troye et al. 2025) and reduced externalizing behaviours (Yildirim and Roopnarine 2015).

### Family functioning in children and young people (CYP) with ND-CNVS

In addition to the psychiatric conditions mentioned above, CYP with ND-CNVs are at elevated risk of irritability (Hall et al. 2024), neurological conditions (Cunningham et al. 2020), difficulties with (sensori-)motor coordination (Cunningham et al. 2019, Cunningham et al, 2021), sleep disturbances (Chawner et al. 2023), seizures (Eaton et al. 2019) and physical health conditions (Óskarsdóttir et al. 2023). Caregivers can struggle to access the care they think their CYP needs (Butter et al. 2024; O’Donovan et al. under review). Consequently, looking after CYP with ND-CNVs can place a considerable burden on families and can impact the quality of life and well-being of caregivers (Butter et al. 2024; Niarchou et al. 2023), with implications for family functioning.

Yet, to our knowledge, only a handful of studies have investigated the associations between family functioning and psychiatric outcomes in CYP with ND-CNVs. Those that have been published to date have focussed on a single ND-CNV - deletion of 22q11.2 leading to 22q11.2 deletion syndrome (22q11.2DS) (Bassett et al. 2011). Prinzie et al (2004) linked parental warmth and control to personality traits such as conscientiousness, emotional stability and independence in children with 22q11.2DS, but found no association with IQ. Allen et al (2014) found that physical punishment, parental control and family organisation significantly predicted social-behavioural and cognitive outcomes. Both studies relied on relatively small samples (n=48 for both studies) and examined behaviours rather than neuropsychiatric outcomes. Furthermore, we are not aware of any studies that have examined differences in caregiver–child relationships between CYP with ND-CNVs and their unaffected siblings, nor whether CYP with ND-CNVs are more vulnerable to the adverse effects of poorer family functioning. Finally, although CYP with inherited ND-CNVs have been shown to exhibit greater behavioural difficulties (Wolstencroft et al. 2022), it remains unclear whether family functioning differs between families with and without an affected parent.

Previous research has underscored the role of other family environmental factors—such as socioeconomic status (SES)—in shaping clinical outcomes in individuals with rare genetic variants, indicating it is important to take these into account when studying family functioning. For example, associations have been reported in CYP with 22q11.2DS between lower SES and both lower social competency and more frequent behavioural difficulties (Shashi et al. 2010; Shashi et al. 2012). Furthermore, a study of a large cohort of CYP with a range of genetic variants found more behavioural difficulties in those growing up in more socioeconomically deprived areas (Wolstencroft et al. 2022).

The current work aims to address the gaps in the literature by investigating both dyadic and systemic measures of family functioning in 553 families with a CYP with a ND-CNV and their unaffected siblings (controls) and evaluating the associations with CYPs psychiatric health and intellectual function.

Specifically, our aims were to:

1. Compare family functioning - as indexed by overall family climate (cohesion/conflict) and caregiver-CYP relationships (warmth/hostility) - in families affected by ND-CNVs with published data on neurotypical families and families under strain (e.g., with depressed individuals, children in crisis, psychiatric patients and alcohol use problems).

Furthermore, in the families affected by ND-CNV, to investigate whether:

2. Caregiver-CYP relationships (warmth/hostility) differ between CYP with ND-CNV and sibling controls.
3. Family functioning is linked with psychiatric outcomes and intellectual functioning in CYP with ND-CNV
4. Family SES (income and maternal education) and inheritance status of the ND-CNV (inherited versus *de novo*) contributes to explaining these relationships
5. The associations between family functioning and psychiatric outcomes and intellectual functioning differ for CYP with ND-CNV and sibling controls

## Methodology

### Sample

553 families with 598 CYP with one (or more) of 15 pathogenic ND-CNVs (Supplementary Table 1) and 222 sibling controls without a ND-CNV were recruited through the Cardiff rarE genetiC variant researcH prOgramme (ECHO) and the IMAGINE-ID (Intellectual Disability & Mental Health: Assessing the Genomic Impact on Neurodevelopment) consortium. via UK Medical Genetics clinics, charities supporting chromosomal conditions (Unique, MaxAppeal) and word-of-mouth (see Chawner et al 2019, for cohort description). A sibling without pathogenic ND-CNV (control) and closest in age to the index child was also invited to take part. The sociodemographic characteristics of the sample are described in Table 1.

**Table 1.** Family background, demographics and family functioning of the families and CYP. Group differences (ND-CNV vs sibling controls) in demographic information were assessed using chi-square tests for sex and an independent t-test for age. Family functioning includes overall family climate (cohesion and conflict) and caregiver-CYP warmth and hostility for individual relationships with ND-CNV CYP and sibling controls. Family climate in families affected by ND-CNV was compared to published means from neurotypical families, families under strain and 22q11.2DS families using one-sample t-tests. Group differences (ND-CNV vs sibling controls) were assessed using mixed effect linear regression for caregiver-CYP relationships, and comparison to published data from neurotypical families was assessed using one-sample t-tests. Significant results are highlighted in bold text.

*Family Background (n = 553 families)*
| **Maternal Education (n = 545)** |  |
| --- | --- |
| No school leaving exams | 36 (6.6%) |
| Low (O-levels, GCSEs) | 118 (21.7%) |
| Middle (A-levels, highers, vocational training) | 188 (34.5%) |
| High (university/post-graduate degree) | 203 (37.2%) |
| **Approximate Family Income (n = 516)** |  |
| ≤£ 19,999 | 144 (27.9%) |
| £ 20 000–£ 39 999 | 169 (32.8%) |
| £ 40 000–£ 59 999 | 108 (20.9%) |
| £ 60 000+ | 95 (18.4%) |
| **Maternal Ethnicity (n = 550)** |  |
| European | 513 (93.3%) |
| Other | 37 (6.7%) |

*Demographics of children and young people*
|  | **ND-CNV**<br>**(n = 598)** | **Sibling controls**<br>**(n = 222)** | **Statistic** |
| --- | --- | --- | --- |
| Female (%) | 239 (40.0%) | 114 (51.4%) | $\chi^2 = 211.92, df = 3, p < 2.2 \times 10^{-16}$ |
| Male (%) | 359 (60.0%) | 108 (48.6%) |  |
| Age (years)<br>Mean ± SD | 11.27 ± 2.97 | 10.18 ± 3.16 | $t = 4.62, p = 5.15 \times 10^{-06}$ |

*Family Functioning: Family Climate <sup>a</sup>*
|  | **Families**<br>**affected by**<br>**ND-CNV**<br>**(n = 553)** | **Neurotypical**<br>**families <sup>b</sup>**<br>**(n = 1125)** | **Families**<br>**under**<br>**strain<sup>b</sup>**<br>**(n = 288)** | **22q11.2DS**<br>**families <sup>c</sup>**<br>**(n = 48)** | **Neurotypical**<br>**families** | **Statistic**<br>**ND-CNV vs**<br>**Families**<br>**under**<br>**strain** | **22q11.2DS**<br>**families** |
| --- | --- | --- | --- | --- | --- | --- | --- |
| Cohesion<br>Mean ± SD | 7.07 ± 0.92 | 6.73 ± 1.47 | 5.25 ± 2.13 | 7.22 ± 2.25 | $t = 8.24, df = 552, p < 2.20 \times 10^{-16}$ | $t = 46.48, df = 552, p < 2.20 \times 10^{-16}$ | $t = -3.64, df = 552, p < 2.94 \times 10^{-04}$ |
| Conflict<br>Mean ± SD | 4.60 ± 1.17 | 3.18 ± 1.91 | 4.02 ± 2.07 | 2.14 ± 1.97 | $t = 28.50, df = 552, p < 2.20 \times 10^{-16}$ | $t = 11.65, df = 552, p < 2.20 \times 10^{-16}$ | $t = 49.37, df = 552, p < 2.20 \times 10^{-16}$ |

*Family Functioning: Caregiver-CYP relationship*
|  | **ND-CNV**<br>**(n = 598)** | **Sibling**<br>**controls**<br>**(n = 222)** | **Neurotypical**<br>**families**<br>**Mean (n)** | **ND-CNV vs sibling**<br>**control** | **Statistic**<br>**ND-CNV vs**<br>**Neurotypical**<br>**mean** | **Sibling controls**<br>**vs Neurotypical**<br>**mean** |
| --- | --- | --- | --- | --- | --- | --- |
| Warmth<br>Mean ± SD | 5.54 ± 0.55 | 5.48 ± 0.57 | 5.7 (96) <sup>d</sup> | $\beta = 0.07, CI = -0.04 - 0.18, p = 1.88 \times 10^{-01}$ | $t = -7.16, df = 597, p = 2.39 \times 10^{-12}$ | $t = -5.65, df = 221, p = 4.82 \times 10^{-8}$ |
| Hostility<br>Mean ± SD | 1.61 ± 0.55 | 1.66 ± 0.53 | 1.69 (740) <sup>e</sup> | $\beta = -0.13, CI = -0.25 - 0.00, p = 4.40 \times 10^{-02}$ | $t = -3.97, df = 597, p = 7.91 \times 10^{-5}$ | $t = -1.06, df = 221, p = 2.92 \times 10^{-1}$ |

a. This measure is an assessment of family climate at the level of the family unit, thus separate values are not given for ND-CNVs and sibling controls. b. Means reported in Moos & Moos (2009). c. Means reported in Allen et al (2014). d. Means imputed from Paine et al (2021). e. Means imputed from Harold & Conger (1997)

Presence or absence of these ND-CNVs was confirmed from Medical Genetics clinics reports and/or biological samples from the CYP and their parents. Biological samples were screened for 54 CNVs considered to be recurrent, pathogenic, or likely pathogenic, as per the American College of Medical Genetics and Genomics guidelines (Richards et al. 2015) and associated with high risk of neurodevelopmental conditions (Kendall et al. 2017) using the Cardiff University Centre for Neuropsychiatric Genetics and Genomics (CNGG) DRAGON pipeline (Lynham et al. 2023). In 197 families neither parent had a ND-CNV, in 95 families at least one parent had a ND-CNV, and the genetic status of both parents was unknown in 261 families.

Written consent was given by the primary caregiver and consent/assent by the CYP as applicable. The authors assert that all procedures comply with ethical standards of the relevant national and institutional committees on human experimentation and with the Helsinki Declaration of 1975, as revised in 2008. All procedures were approved by the appropriate university and National Health Service (NHS) ethics and research and development committees; the NHS London Queen Square (14/LO/1069), South East Wales (09/WSE04/22), East Midlands - Leicester Central (19/EM/0287), London – Fulham (24/LO/0689), and London South East (20/LO/1271).

### Measures

#### Family Functioning

##### Family climate

The Family Environment Scale (FES) measures the overall social and environmental characteristics of families (Moos and Moos 2009) and is therefore completed once by the caregiver capturing information for both CYP with ND-CNV and siblings. It measures two constructs: cohesion and conflict, each comprising 9 items. Family cohesion reflects commitment, helpfulness and support between family members (example statement: ‘There is a feeling of togetherness in our family’), whereas family conflict assesses open expression of anger, aggression and conflict between family members (example statement:’ Family members sometimes get so angry they throw things’). Items are rated on a 4-point Likert scale ranging from 1 (*Strongly disagree*) to 4 (*Strongly agree*), some of which are reverse coded before averaging across items. Ratings are given per family (range 0 – 9). Higher scores reflect a higher level of the construct (conflict or cohesion). Moos and Moos (2009) report good internal consistency and test-retest reliability for the subscales.

##### Caregiver-CYP relationship

Warmth and hostility in the relationship between the caregiver and each participating CYP individually (both with ND-CNV and sibling controls) was assessed using the Behavioral Affect Rating Scale (BARS, Conger 1989), which queries how often caregivers acted in certain ways toward their child during the last month. The warmth subscale (Paine et al. 2021) comprises six items (example statement: ‘how often do you act loving and affectionate towards them?’) and the hostility subscale (Harold and Conger 1997) four items (example statement: ‘how often do you get angry with them?’). Responses are made on a seven-point Likert scale ranging from 1 (*always*) to 7 (*never*). All items were reverse coded, so that higher scores on the hostility subscale indicate a more hostile relationship, while higher scores on the warmth subscale are indicative of a warmer relationship and a mean score for each subscale was subsequently calculated (warmth range 0-6, hostility range 0-4). The BARS subscales have demonstrated good reliability and internal consistency (Harold and Conger 1997).

#### Assessment of neuropsychiatric and cognitive phenotypes Psychopathology

The child and adolescent psychiatric assessment (CAPA) is an in-depth, semi-structured research diagnostic psychiatric interview (Angold and Costello 2000), which was carried out by experienced research psychologists with the primary caregiver. A research diagnosis based on the DSM-V was obtained through consensus meetings led by a child and adolescent psychiatrist.

We used the CAPA to establish both externalising and internalising symptoms and diagnoses. Externalising symptoms and diagnoses included ADHD and ODD. Internalising symptoms and diagnoses included anxiety (e.g., generalised anxiety disorder, social phobia, specific phobia, separation anxiety, panic disorder with and without agoraphobia, and panic disorder), mood disorder (depression, mania or hypomania) and obsessive-compulsive disorder (OCD). We did not consider any symptoms or diagnoses to be mutually exclusive.

#### Cognition

The young person’s intelligence quotient (IQ) was assessed using the Wechsler Abbreviated Scale of Intelligence (WASI), from which full-scale IQ (FSIQ), verbal IQ (VIQ), and performance IQ (PIQ) composite scores were derived.

### Analysis

The data were analysed using R version 4.4.2.

#### Aim 1

To investigate overall family climate we calculated the mean raw values for the two FES subscales (cohesion and conflict) across our sample and conducted comparisons to published norms from neurotypical populations (Moos and Moos 2009), families under strain (including families with depressed individuals, children in crisis, psychiatric patients and substance use, Moos and Moos 2009), and published data from 22q11.2DS families (Allen et al. 2014) using one-sample t-tests.

As a subset of families were affected by 22q11.2DS (*n* = 171), secondary analyses compared cohesion and conflict in these families to published data from 22q11.2DS families (Allen et al., 2014).

We calculated mean BARS scores to evaluate warmth and hostility in the relationships between caregivers and CYP with ND-CNV and controls, and conducted comparisons to published data imputed from neurotypical populations (Harold and Conger 1997; Paine et al. 2021) using one-sample t-tests.

We conducted secondary analyses comparing the mean family functioning measures in families with (n=95) and without an affected parent (n=197) using independent t-tests. Of the families with an affected parent, 63 comprised families with an affected mother, 30 with an affected father, and 2 families in which both parents were affected.

#### Aim 2

To compare caregiver-CYP relationships in CYP with ND-CNVs and controls, we first transformed the raw family functioning measures using the Tukey Ladder of Powers transformation to make the data fit the normal distribution as closely as possible. We then standardised these transformed scores into Z scores using the mean and SD of the whole sample as reference. We ran mixed effects linear regression models examining the association between ND-CNV status and caregiver-CYP measure warmth and hostility separately, including age and sex as covariates and family membership as a random effect - to take into account that CYP with ND-CNV and siblings from the same family are more likely to share genetic and other family background factors (e.g. scaled caregiver-CYP measure ∼ ND-CNV status + age + sex + (1| family membership)).

#### Aim 3

To compare the relationship between family functioning and psychiatric and cognitive outcomes in CYP with ND-CNVs, we transformed the psychiatric symptom scores using the Tukey Ladder of Powers and then transformed these scores into Z scores using the mean and SD of the whole sample as reference. All subsequent models were run on these transformed family functioning and psychiatric scores.

Separate linear regression models (e.g. scaled symptom outcome∼ scaled family functioning measure + age + sex) were run for each family functioning measure (cohesion, conflict, warmth and hostility) with symptoms related to externalizing conditions (e.g. ODD and ADHD), and internalizing symptoms (e.g., mood, anxiety and OCD), and cognition (FSIQ, VIQ and PIQ) in CYP with ND-CNVs only.

Of the 553 families in the study, 536 (96.9%) reports were from mothers, 14 (2.5%) from fathers and 3 (0.5%) from other caregivers (e.g. grandparents). We conducted a sensitivity analysis rerunning the models involving caregiver-CYP warmth and hostility using mothers’ reports only (n= 536 families, with 581 CYP with ND-CNVs). Additional sensitivity analyses reran the models for family functioning measures with parental affected status as a covariate.

#### Aim 4

To examine the contribution of SES to family functioning, we first conducted separate linear regression models (e.g. Family functioning measure ∼ SES measure + age + sex) with each family functioning measure (cohesion, conflict, warmth and hostility) in a subset of families for whom this data was available (Income: n= 516 (93.3%), Education: n = 545 (98.6%)).

Subsequent analyses included family income and maternal education as covariates in the Aim 3 regression models to test whether SES contributes to explaining the relationships between family functioning and psychiatric outcomes.

#### Aim 5

To evaluate whether the association between family functioning and psychiatric and cognitive outcomes differs between CYP with ND-CNV and controls we included an interaction term between the family functioning measures and ND-CNV status in models in which we observed a significant main effect of family functioning in Aim 4 (e.g. ODD ∼ CNV status + cohesion + CNV status * cohesion + age + sex + income + education (1| family membership)).

False discovery: We applied Benjamini–Hochberg (BH) false discovery rate (FDR) corrections (Benjamini and Hochberg 1995) to the primary analyses to control for false discovery due to multiple statistical tests. Because the pre-specified aims addressed conceptually distinct questions, each aim was treated as a separate family of analyses. Within Aims 3 and 4, corrections were applied across the (psychiatric and cognitive) symptom domains (n=8) and family functioning measures (n=4) evaluated (24 tests). Interaction terms (Aim 5) were considered secondary exploratory analyses and were not included in the primary FDR correction (Benjamini and Yekutieli 2001).

## Results

### Family functioning in families affected by ND-CNV

Families affected by ND-CNVs had increased levels of cohesion compared to both neurotypical families as well as families under strain, but reduced levels compared to families with a CYP with 22q11.2DS. Families affected by ND-CNVs also had increased levels of conflict compared to both neurotypical families, families under strain and families with a CYP with 22q11.2DS, see Table 1 and Figure 1. Comparisons limited to the families affected by 22q11.2DS showed that cohesion was similar to published data from 22q11.2DS families, but family conflict was greater, see Supplementary Table 3.

**Figure 1.**
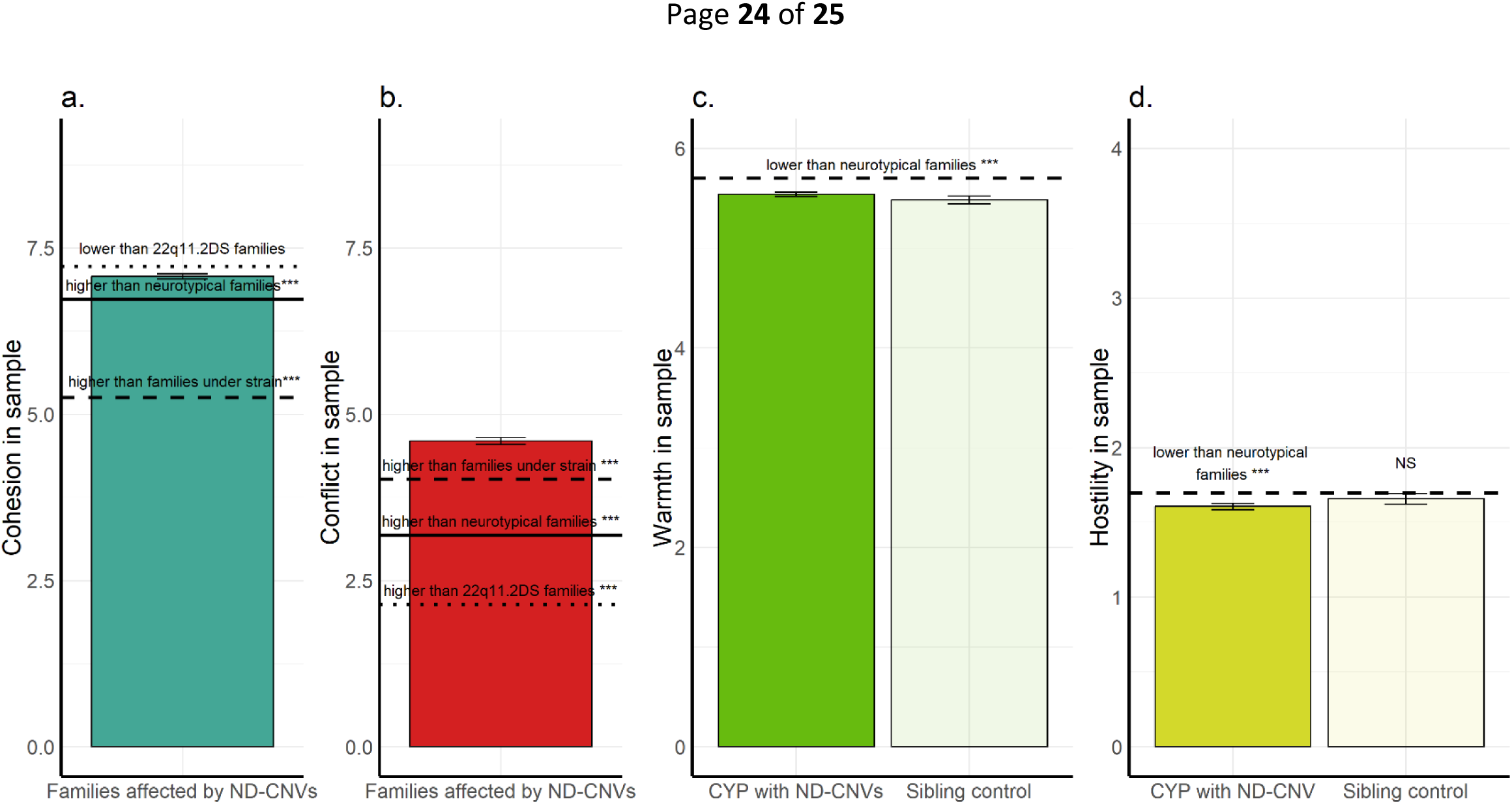
Family functioning in families affected by ND-CNV compared to published data from neurotypical families, families under strain (Moos and Moos, 2009) and 22q11.2DS families (Allen et al., 2014). Family climate: a) cohesion and b) conflict in families affected by ND-CNV. Solid black lines represent data from neurotypical families. Black dashed lines represent data derived from families under strain including with depressed individuals, children in crisis, psychiatric patients and alcohol use problems. Black dotted lines represent data from 22q11.2DS families. Caregiver-CYP c) warmth and d) hostility. Black dashed lines represent published data from neurotypical families for warmth and hostility. *** p < 0.001 NS = non-significant. Demographic information for these samples, where available, is provided in Supplementary Table 2. Cohesion and conflict (a and b) are family level measures, whereas caregiver-CYP warmth and hostility (c and d) are given for each relationship with a CYP.

Levels of warmth reported for CYP with ND-CNVs and controls were reduced compared to neurotypical families. Levels of hostility were reduced for CYP with ND-CNV, but comparable with published data for controls, see Table 1 and Figure 1.

We found increased family conflict and decreased cohesion in families comprised of a parent affected by ND-CNV compared to families with no affected parents. Both families with an affected parent(s) and families where both parents are unaffected by ND-CNV showed increased cohesion and increased conflict compared to neurotypical families. No differences were observed in caregiver-CYP relationships, see Supplementary Table 4.

### Reduced caregiver-CYP hostility to CYP affected by ND-CNV compared to unaffected siblings

ND-CNV was associated with caregiver-CYP hostility, with less caregiver-CYP hostility reported toward CYP with an ND-CNV. No differences between caregiver-CYP relationship warmth were observed between CYP with ND-CNVs compared to sibling controls, see Table 1.

### Family functioning in CYP affected by ND-CNV is associated with psychiatric outcomes

Psychiatric and cognitive outcomes are summarised in Table 2. Compared with controls, CYP with ND-CNVs showed higher levels of internalising (OCD, anxiety, and mood) and externalising (ADHD and ODD) symptoms and diagnoses, as well as lower IQ. Table 3 presents regression analyses examining associations between family functioning and CYP psychiatric and cognitive outcomes, controlling for age, sex, and SES, with uncorrected and corrected *p*-values reported.

**Table 2.**
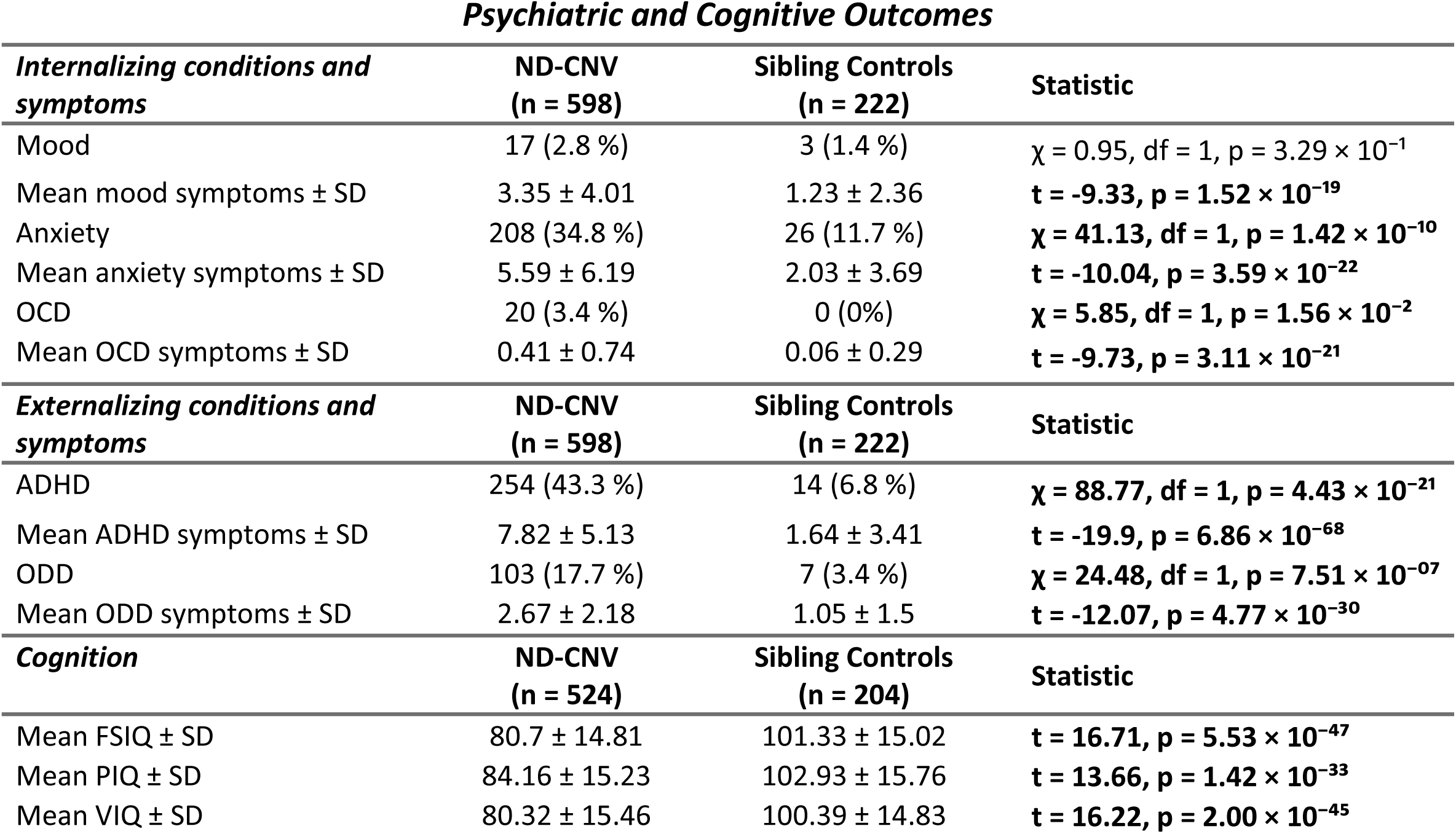
Psychiatric diagnosis and symptoms and cognition in CYP with ND-CNVs and sibling controls. Number and percentage of young people meeting criteria for psychiatric diagnoses, in addition to symptom counts, and cognitive ability. Diagnoses include internalizing conditions: Mood disorders (including depression and mania), Any anxiety diagnosis (including generalized anxiety disorder, social phobia, specific phobia, separation anxiety, panic disorder with and without agoraphobia, and panic disorder), and Obsessive Compulsive Disorder (OCD), and externalizing conditions: Attention Deficit Hyperactivity Disorder (ADHD) and Oppositional Defiant Disorder (ODD). Cognitive ability is summarized for Full Scale IQ (FSIQ), Verbal IQ (VIQ), and Performance IQ (PIQ). Group differences (ND-CNV vs sibling controls) were assessed using chi-square tests for categorical variables and independent t-tests for continuous variables. Significant differences are highlighted with bold text.

**Table 3.**
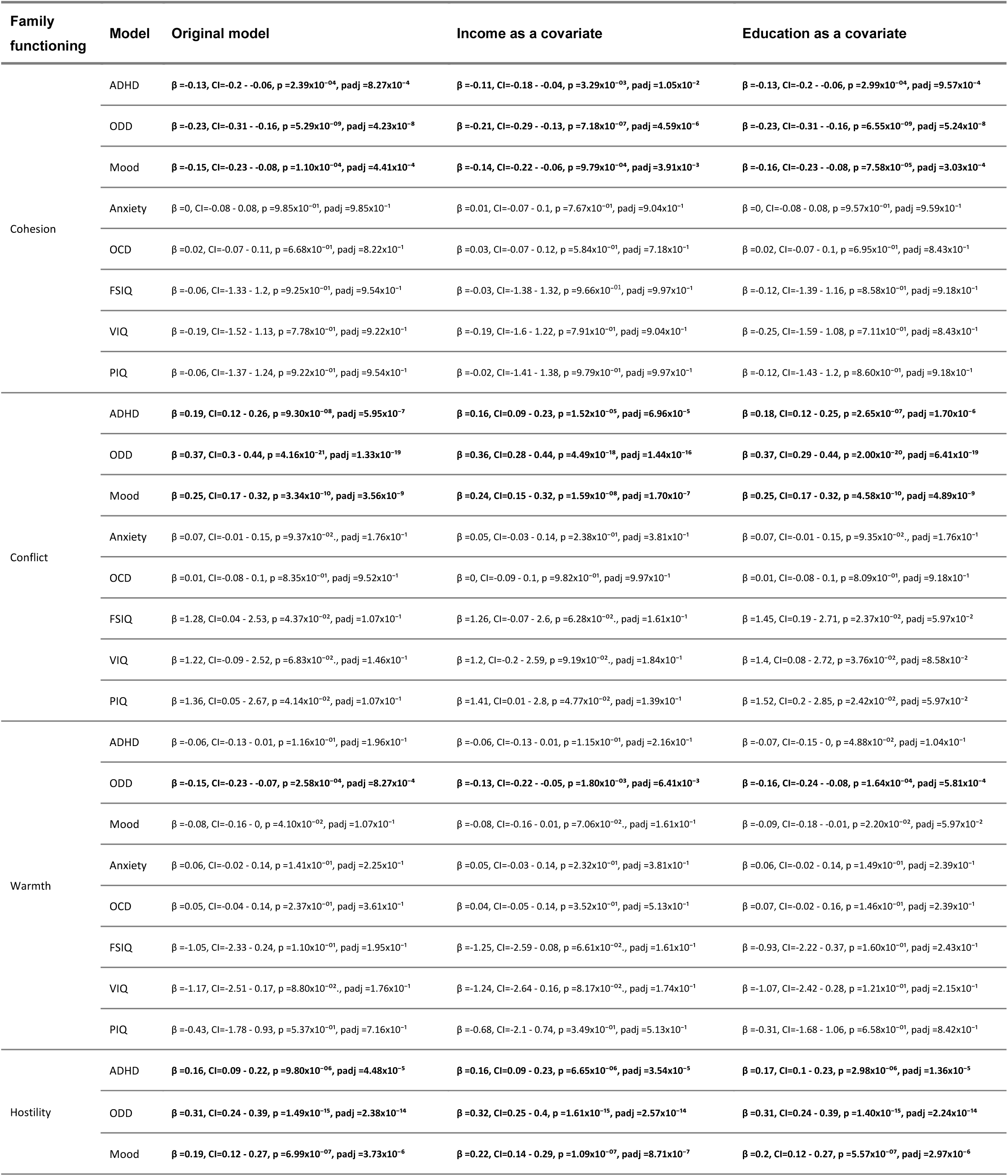
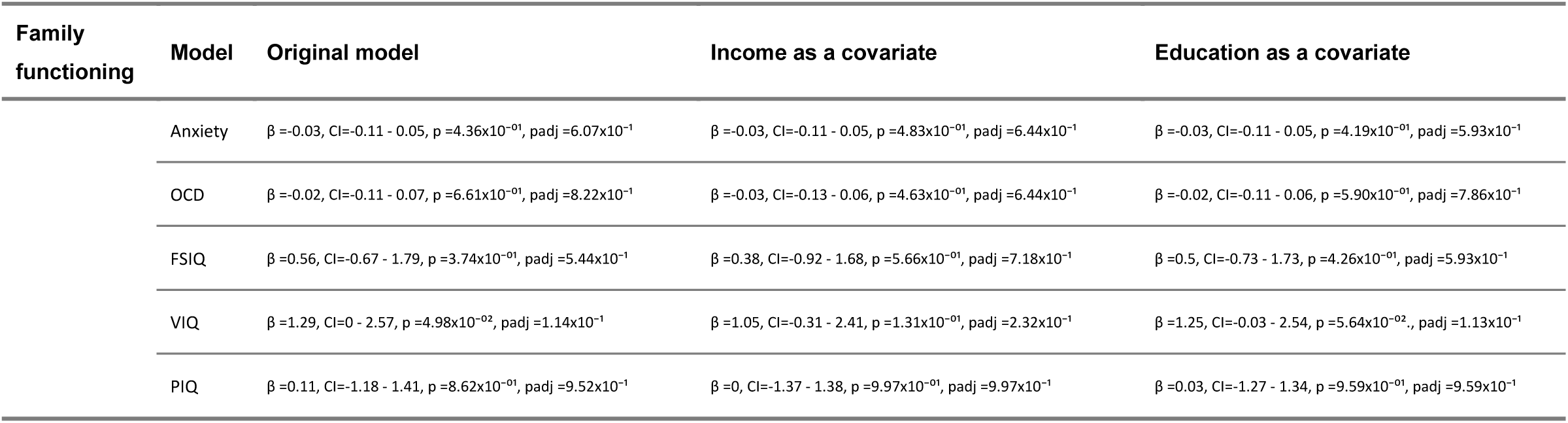
Regression models assessing the associations between family functioning and psychiatric and cognitive outcomes in CYP with ND-CNV. Main effects of family functioning measures while controlling age and sex assigned at birth (Aim 3), and with family income and maternal education added as covariates (Aim 4). Models were conducted with normalized z-scores for both family functioning measures and symptoms counts for psychiatric outcomes (ADHD, ODD, Mood, Anxiety, and OCD), and age-standardised scores for cognitive outcomes (FSIQ, VIQ and PIQ). Padj = p-value corrected for multiple comparisons (corrected across 24 comparisons within each set of models using Benjamini–Hochberg). Associations that remained significant following correction are highlighted in bold text.

In CYP with ND-CNV, higher levels of family cohesion were associated with lower externalising symptoms: ADHD (β = -0.13) and lower ODD (β = -0.23), as well as reductions in internalising symptoms related to mood disorders (β = -0.15). Increased levels of family conflict were associated with higher externalising symptoms (ADHD (β = 0.19) and ODD (β = 0.37)), as well as more mood disorder symptoms (β = 0.25).

Increased caregiver-CYP warmth was associated with reductions in ODD (β = -0.15) as well as mood disorder symptoms (β = -0.08), although the latter did not survive FDR correction. Higher levels of caregiver-CYP hostility were associated with increased ADHD (β = 0.16),

ODD (β = 0.31) and mood disorder symptoms (β = 0.19). No associations were found between family cohesion, conflict, warmth or hostility and CYP anxiety or OCD symptoms.

Increased family conflict was associated with a small increase in both FSIQ (β = 1.28) and PIQ (β = 1.36). Caregiver-CYP hostility was associated with VIQ only (β = 1.29). No other associations were found between any measures of family functioning and IQ, and none of these associations survived correction for multiple testing, see Table 3.

The associations between family functioning and mental health outcomes remained similar when controlling for parental ND-CNV status, and the associations with caregiver-CYP relationships were similar when restricted to maternal caregiver relationships (Supplementary Tables 5 and 6, respectively).

### Family income and maternal education are associated with family functioning but do not contribute to associations between family functioning and psychiatric symptoms

Family income was associated with increased family cohesion (β = 0.16) and reduced conflict (β = -0.23). Maternal education was associated with reduced family conflict (β = -0.42), see Supplementary Table 7.

The associations between family climate or caregiver-CYP relationship and psychiatric outcomes and intellectual functioning remained broadly similar when controlling for family income and maternal education (Table 3).

### ND-CNV * family climate interactions and psychiatric symptoms

We subsequently tested whether the associations between family functioning and ADHD, ODD and mood symptoms differed between CYP with ND-CNVs and controls. An interaction between family cohesion and ODD symptoms was observed (β = -0.14, Figure2a): showing that increased family cohesion was associated with a larger decrease in ODD symptoms in CYP with ND-CNVs compared to controls. Furthermore, increased conflict was associated with more ODD symptoms in CYP with ND-CNVs compared to sibling controls (β = 0.18, Figure 2b). Finally, increased caregiver-CYP hostility was associated with a larger increase in mood disorder symptoms in CYP with ND-CNVs than sibling controls (β = 0.15, Figure2c). See also Supplementary Table 8.

**Figure 2.**
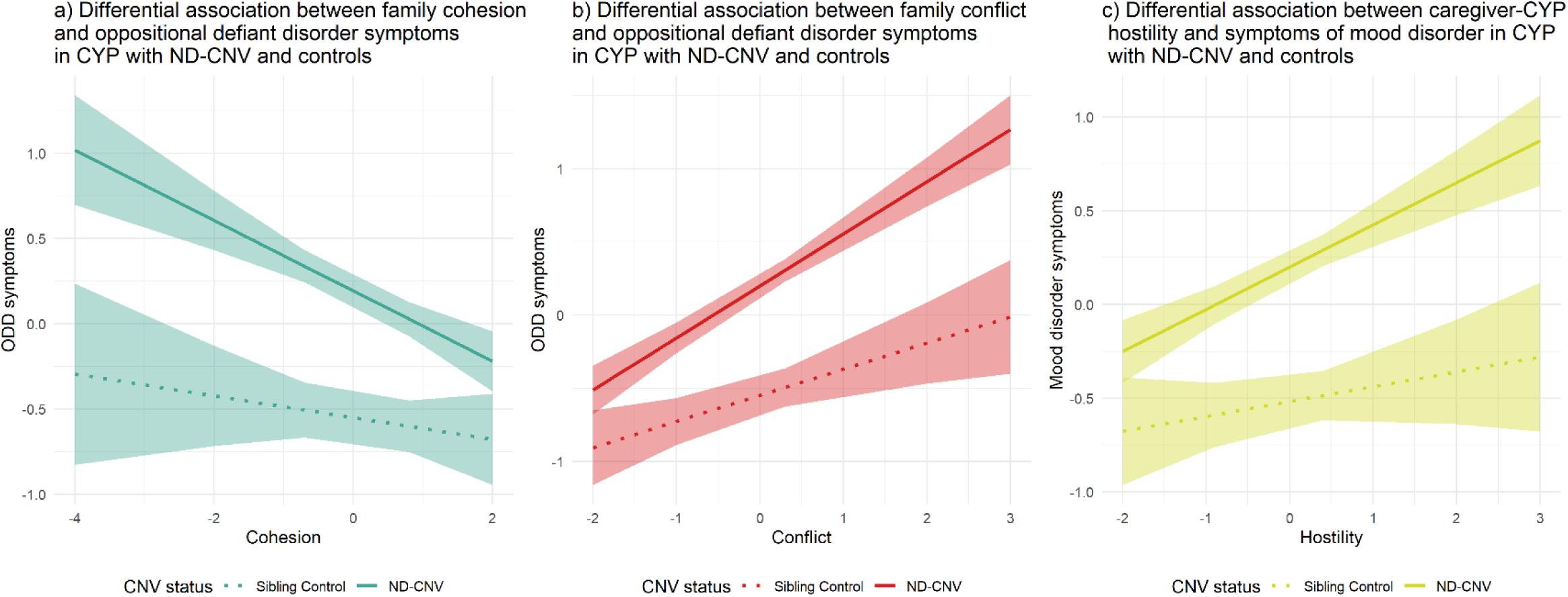
Plots of the significant family functioning by ND-CNV status interactions. Differential associations between a) family cohesion and oppositional defiant disorder symptoms, b) family conflict and oppositional defiant disorder symptoms, and c) caregiver-CYP hostility and mood disorder symptoms, in CYP with ND-CNV and controls. X-axis denotes family functioning measure (normalized z-score); Y-axis denotes psychiatric symptom counts (normalized z-score) for ODD (a and b) and mood disorder (c); legend denotes CNV status (ND-CNV - solid line, sibling control - dotted line); shaded areas represent 95% confidence intervals.

## Discussion

The sources of variability in phenotypic outcomes among CYP with ND-CNVs (e.g., Chawner et al. 2019; Jacquemont et al. 2022) remain poorly understood, although environmental factors are likely to contribute. To our knowledge, this is the first study to investigate the relationship between family functioning and neurodevelopmental and mental health outcomes in CYP with a range of rare genetic mutations that are associated with intellectual and developmental disabilities. We examined a cohort of 820 CYP, including 598 with one of 16 ND-CNVs and 222 unaffected siblings. Compared with published data, families affected by ND-CNVs showed higher levels of both family cohesion and conflict than neurotypical families and families experiencing strain. Caregiver–CYP warmth was lower in ND-CNV families relative to neurotypical norms, whereas caregiver–CYP hostility was reduced for CYP with ND-CNVs but not for siblings. Family climate and caregiver–CYP relationship quality were associated with both internalising (mood disorder) and externalising (ADHD and ODD) symptoms. Although maternal education and family income were associated with family functioning, these factors did not account for the associations between family functioning and psychiatric symptoms. Interaction analyses indicated that higher family cohesion was associated with lower ODD symptoms in CYP with ND-CNVs relative to controls, whereas higher family conflict was associated with higher ODD symptoms. Greater caregiver–CYP hostility was additionally associated with increased mood disorder symptoms in CYP with ND-CNVs. No associations were observed between family functioning and IQ.

We observed a distinct pattern of family functioning relative to published data from families with neurotypical children, marked by lower caregiver warmth to both CYP with and without ND-CNV, reduced hostility to CYP with ND-CNV, and at the family level, reduced cohesion, and higher conflict. The pattern also diverged from published data on families under strain. Specifically, cohesion and conflict were both elevated in families affected by ND-CNV. Many factors may combine to impact family functioning, and their inter-relationships are often complicated. There is evidence for bidirectional relationships between child mental health symptoms and family functioning with evidence for both child effects on parenting and family environment/parental style influences on the child (e.g., Hickey et al. 2020). Moreover, family stressors, including negative life events and parental psychological distress, have been linked to decreased parental warmth and increased child externalizing behaviours (Park and Dotterer 2018).

In families affected by ND-CNV, the stressors and burden which come with living with a CYP who may have additional physical and mental health needs are likely to impact caregiver-child interactions, and family functioning more widely. Parents with CYP with a ND-CNV may experience poorer social support (Currie and Szabo 2020) and more mental health issues (Niarchou et al. 2023). There may also be financial strains because of the time commitment involved with caring for CYP with additional needs (Butter et al. 2024; van den Bree et al. 2013). Interestingly, family functioning in our sample was found to be associated with maternal education and family income. Family income was associated with increased cohesion and reduced conflict, whereas maternal education was linked to reduced family conflict. Thus, increased education and income may interact with family functioning by making it easier to access support, as well as capacity to advocate for their child’s needs.

Families affected by 22q11.2DS in our sample reported higher conflict than those in a smaller U.S. sample (Allen et al., 2014). Given the lower socioeconomic status of the present sample (Supplementary Table 2) and observed associations between income and conflict, socioeconomic differences may to an extent explain this variation.

We found decreased family cohesion and increased conflict in families with affected parent(s) compared to families where both parents are unaffected. However, in families with unaffected parents we still observed increased conflict and increased cohesion compared to published data from neurotypical families. The effects of caring for CYP with ND-CNVs are likely exacerbated in families in which the ND-CNV is inherited (Wolstencroft et al. 2022), possibly as an affected parent might find it harder to care and advocate for a child with multiple needs. Further studies examining the nature and interplay between family environment and the impact of shared genetics (e.g. ND-CNV inheritance and polygenic risk for mental health outcomes) are required to understand the full impact that family functioning can have on the aetiology and expression of mental health outcomes.

This work extends previous results in 22q11.2DS to a broad range of ND-CNVs and neuropsychiatric outcomes: we found in both CYP with ND-CNV and controls family conflict and caregiver-CYP hostility were associated with an increase in mood disorder symptoms. Conversely, family cohesion and caregiver warmth were associated with reduced mood disorder symptoms in our sample. This is in support of previous studies that show that low family cohesion, high conflict, and poor parental support are associated with increased depressive symptoms (Yu et al. 2015). We also found that ADHD and ODD symptoms were associated with family functioning. These findings are in line with previous research showing that higher family conflict and caregiver hostility are linked to higher levels of externalising symptoms in children (McKinney and Renk 2011); and that caregiver warmth (Pinquart 2017) and family cohesion are associated with reduced externalising problems (Sbicigo and Dell’Aglio 2013). Consistent with previous reports in 22q11.2DS (Prinzie et., al 2004), we found no associations between family functioning and IQ.

Our interaction analyses indicated that associations between family functioning and psychiatric symptoms differed between CYP with ND-CNVs and sibling controls. Associations between negative family functioning and psychiatric outcomes (conflict and ODD symptoms and hostility and mood symptoms), as well as between family cohesion and ODD symptoms, were stronger in CYP with ND-CNVs. These interactions are likely to reflect the bidirectional interplay between systemic family functioning and psychiatric symptoms in CYP with rare genetic mutations associated with intellectual and developmental disabilities. Importantly, although lower socioeconomic status has been associated with clinical outcomes in 22q11.2DS (Shashi et al., 2010, 2012), the associations and interactions between family functioning and psychiatric symptoms remained after controlling for maternal education and family income.

This study provides the first evidence of the association between family functioning and psychiatric outcomes in CYP with ND-CNVs. Raising a CYP with neurodevelopmental, and often additional complex, needs can be challenging (Butter et al. 2024; van den Bree et al. 2013). Moreover, mothers of CYP with ND-CNV have been shown to be at increased risk of psychiatric difficulties (Niarchou et al. 2023). Our findings highlight the need to include family functioning measures in both research and clinical practice to inform risk for psychiatric outcomes in genetically vulnerable populations. Families and CYP with ND-CNVs may benefit from interventions, such as multidimensional family therapy (e.g., Hogue et al. 2006), and support targeted at enhancing cohesion and reducing conflict and caregiver-CYP hostility to help mitigate psychiatric risk. Timely access to psychiatric support for the CYP - as well as for parents, where needed - may also help family functioning and have positive effects for the entire family.

## Data Availability

All data produced in the present study are available upon reasonable request to the authors

## Abbreviations

22q11.2DS: 22q11.2 deletion syndrome
ADHD: attention deficit hyperactivity disorder
BARS: Behavioral Affect Rating Scale (Conger, 1989)
BH: Benjamini-Hochburg
CAPA: child and adolescent psychiatric assessment (Angold & Costello, 2000)
CNV: copy number variant
CYP: children and young people
ECHO: Cardiff rarE genetiC variant researcH prOgramme
FDR: False Discovery Rate
FES: Family Environment Scale (Moos & Moos, 2009)
FSIQ: full scale intelligence quotient
IQ: Intelligence Quotient
ND-CNV: neurodevelopmental copy number variants
OCD: obsessive-compulsive disorder
ODD: oppositional defiant disorder
PIQ: performance intelligence quotient
SES: socioeconomic status
VIQ: verbal intelligence quotient
WASI: Wechsler Abbreviated Scale of Intelligence

## Acknowledgements

We are extremely grateful to all the young people and their families, who took part in the Cardiff rarE genetiC variant researcH prOgramme (ECHO) and Intellectual Disability and Mental Health: Assessing the Genomic Impact on Neurodevelopment (IMAGINE-ID) studies. We thank the UK National Health Service (NHS) medical genetic clinics for their support, and charities Unique and Max Appeal for their support. We thank the core laboratory team of the Division of Psychological Medicine and Clinical Neurosciences Laboratory for DNA sample management and genotyping and thank Alexandra Evans and Nabila Ali for their support with interpreting genotype information. We would like to extend thanks to the ECHO field team over the years: Matthew Sopp, Alice Robinson, Sinéad Ray, Nicola Lewis, Sarah Law, Sophie Andrews, Aimée Challenger, Poppy Sloane, Alice Walsh, Keziah Fish, Amy Ilsley, Stephen Naughton, Rachel Tompkins, Ciara Walker, Nadia Pantouw, Samantha Bowen, Hannah Pendlebury, Chloe Sheldon, Emily Green, Umaya Prasad, Joshua Roberts, Jessica Townsend, Beth Hughes, Rachael Adams, Alister Baird, Hayley Moulding, Sinead Morrison, Adam Cunningham, Ffion Evans, Jacqueline Smith, Holly Howe, Lauren Benger, Hannah Tyson, Claudia Evison, Georgia Evans, Caitlin Goldie, Charlotte Butter, Jack Griffiths, Ioana Filipas, Danielle LeRoux, Shreeya Sivakumar, Sally Morrin, Hannah Thomas, Emily Collins, Elena Bishop, Daniel Brookes and Helena Pumfrey for their roles in data collection. We thank Rhiannon Spence, Tracey Ball and Owen Myers for administrative support. Jeremy Hall gratefully acknowledges the support of the Hodge Foundation and The Waterloo Foundation.

Author contributions: JEH, JHH, JH, MJO, and MBMvdB conceptualised the work. JHH and MBMvdB coordinated the data collection. JEH and JHH prepared the data. JEH analysed the data. JEH, JHH and MBMvdB were involved in the analysis methodology, and interpretation of the data. JEH wrote the original draft of the manuscript. JEH, JHH, JH, MJO, and MBMvdB substantively reviewed and revised the manuscript. The ECHO study was designed by MBMvdB and MJO. The IMAGINE-ID study was designed by MBMvdB, MJO, and JH. MBMvdB, MJO, and JH were involved in funding acquisition.

## IMAGINE-ID Collaborators

David Skuse^1^, **

Jeanne Wolstencroft^1^

Irene O Lee^1^

Samuel Chawner^2^

Jeremy Hall^2,3,4^

Josh Hope-Bell^2^

Michael J Owen^2,3^

Marianne van den Bree^2,3,^ **

F Lucy Raymond^5^

Ramya Srinivasan^6^

** Consortium contact: David Skuse, Paediatric Mental Health Sciences Centre, Great Ormond Street Institute of Child Health, University College London, London, UK; or Marianne van den Bree, Neuroscience and Mental Health Innovation Institute, Division of Psychological Medicine and Clinical Neurosciences, Cardiff University, Cardiff, UK;

^1^ Paediatric Mental Health Sciences Centre, Great Ormond Street Institute of Child Health, University College London, London, UK.

^2^ Centre for Neuropsychiatric Genetics and Genomics, Division of Psychological Medicine and Clinical Neurosciences, Cardiff University, Cardiff, UK.

^3^ Neuroscience and Mental Health Innovation Institute, Division of Psychological Medicine and Clinical Neurosciences, Cardiff University, Cardiff, UK.

^4^ Department of Psychiatry, Warneford Hospital, University of Oxford, UK

^5^ Department of Medical Genetics, University of Cambridge, UK

^6^ Division of Psychiatry, University College London, UK

