## Supplementary Table 1 for "Family functioning and psychiatric outcomes in children and young people with intellectual and developmental disabilities caused by rare genetic mutations"

**Supplementary Table 1. ND-CNVs and sample sizes for participating CYP.**

ND-CNVs were selected for study inclusion based on robustly associated with intellectual disability and neuropsychiatric phenotypes, including schizophrenia and autism spectrum disorder. ND-CNV presence or absence of any pathogenic CNV in controls was confirmed via NHS Medical Genetics records and subsequently, by the laboratory of the Cardiff University Division for Psychological Medicine and Clinical Neurosciences (CU DPMCN), using microarray techniques, referenced to the hg19 genome build.

| **CNV** | **n** | **Critical Region (hg19)** |
| --- | --- | --- |
| Control | 222 | - |
| 22q11.2 deletion | 175 | chr22: 19,037,332-21,466,726 |
| 16p11.2 deletion | 91 | chr16: 29,650,840-30,200,773 |
| 16p11.2 duplication | 51 | chr16: 29,650,840-30,200,773 |
| 22q11.2 duplication | 47 | chr22: 19,037,332-21,466,726 |
| 15q11.2 deletion | 37 | chr15: 22,805,313-23,094,530 |
| 1q21.1 duplication | 36 | chr1: 146,527,987-147,394,444 |
| 15q13.3 deletion | 21 | chr15: 31,080,645-32,462,776 |
| 1q21.1 deletion | 17 | chr1:146,527,987-147,394,444 |
| 15q13.3 duplication | 18 | chr15:31,080,645-32,462,776 |
| 2p16.3 deletion (NRXN1) | 16 | chr2:50145643-51259674 |
| 9q34 deletion (EHMT1) | 11 | chr9:140,513,444-140,730,578 |
| 3q29 deletion | 6 | chr3:195,720,167-197,354,826 |
| 3q29 duplication | 4 | chr3:195,720,167-197,354,826 |
| SHANK3 deletion | 3 | chr22:51113070-51171640 |
| 15q11.2 duplication | 2 | chr15:22,805,313-23,094,530 |
| Multiple CNVs | 63 | - |
