## Supplementary Table 2 for "Family functioning and psychiatric outcomes in children and young people with intellectual and developmental disabilities caused by rare genetic mutations"

**Supplementary Table 2. Details of the families and samples for the ECHO study and previously published data in neurotypical families, families under strain, and 22q11.2DS families.**

| ***Sample*** | ***Number of families*** | ***Country of Origin*** | ***Family and Sample Composition*** |
| --- | --- | --- | --- |
| ECHO cohort  Cardiff rarE genetiC variant researcH prOgramme (ECHO, https://www.cardiff.ac.uk/centre-neuropsychiatric-genetics-genomics/research/themes/developmental-psychiatry/copy-number-variant-research-group). | 553 | UK and Ireland | The child and adolescent participants ranged in age from 6 to 24 years, with an average age of 10.5 years (SD=3.14). The sample was 57% male and predominantly white (93.4%). Income placed the sample within the low-middle socioeconomic stratum. The majority of adults completing the evaluation were mothers. |
| FES norms from neurotypical families  Moos & Moos (2009) | 1125 | United States | Families were recruited from across the United States, including single-parent families, multi-generational families, ethnic minority families and families from various socioeconomic backgrounds. Detailed demographic information for this sample is not provided. |
| FES norms from families under strain  Moos & Moos (2009) | 288 | United States | Families were recruited from across the United States and included 220 families with alcohol use problems, 77 families with psychiatric patients, 161 families with children or adolescents in crisis situations. Detailed demographic information for this sample is not provided. |
| FES data from families affected by 22q11.2 deletion  Allen et al (2014) | 48 | United States | The child and adolescent participants ranged in age from 9 to 18 years, with an average age of 12.46 years (SD=2.07). The sample was 54.2% male and largely white (83.3%). The Hollingshead Two-Factor Index of Social Position (Hollingshead 1957) placed the sample within the middle socioeconomic stratum (Mean =29.3, SD=13.45). The majority of adults completing the parent portion of the evaluation were mothers. |
| Published data on caregiver-child warmth in adoptive families  Paine et al (2021) | 96 | United Kingdom | The child and adolescent participants ranged in age from 1 to 12 years. The sample was 51% male and largely white (99%). The family income and education levels were higher than average. |
| Published data on caregiver-child hostility from neurotypical rural families  Harold & Conger (1997) | 370 | United States | The child and adolescent participants had an average age of 12.7 years old at the first assessment. The sample was 47% male. Families all comprised two-parent households from rural Iowa, with the majority living in small towns. They were in the lower-middle to middle class, average education, but were poorer than average, compared to similar families in the United Stated. Both parents completed the assessment of parent-child relationships. |
