## Supplementary Table 3 for "Family functioning and psychiatric outcomes in children and young people with intellectual and developmental disabilities caused by rare genetic mutations"

**Supplementary Table 3. Family climate in a subset of families affected by 22q11.2DS and comparison to published data.**

Family climate in families affected by 22q11.2DS was compared to published means from 22q11.2DS families using one-sample t-tests. Significant results are highlighted in bold text.

| ***Family Functioning: Family Climate*** | | | |
| --- | --- | --- | --- |
|  | **Families affected by 22q11.2DS**  **(n = 171)** | **22q11.2DS families ^a^**  **published data**  **(n = 48)** | **Statistic** |
| Cohesion  Mean **±** SD | 7.13 ± 0.97 | 7.22 ± 2.25 | t = -1.23, df = 170, p = 2.21 x 10^-1^ |
| Conflict  Mean **±** SD | 4.45 ± 1.13 | 2.14 ± 1.97 | **t = 26.87, df = 170, p < 2.20 x 10^-16^** |

a. Allen et al., 2014
