## Supplementary Table 4 for "Family functioning and psychiatric outcomes in children and young people with intellectual and developmental disabilities caused by rare genetic mutations"

**Supplementary Table 4. Family background and family functioning in families with and without parents affected by ND-CNV**

Family functioning includes overall family climate (cohesion and conflict) and caregiver-CYP warmth and hostility for individual relationships with ND-CNV CYP and sibling controls. Group differences (parent(s) affected by ND-CNV vs unaffected parent families) were using chi-square tests for categorical and independent t-tests for continuous variables. Significant results are highlighted in bold text.

***Family Background (n = 553 families)***

|  | **Parent(s) affected by ND-CNV^a^**  **(n = 95 with 122 CYP with ND-CNV and 21 control sibs)** | **No affected parents**  **(n = 197 with 208 CYP with ND-CNV and 104 control sibs)** | **Parental ND-CNV status unknown**  **(n =261 with 268 CYP with ND-CNV and 97 control sibs))** | **Statistic**  ***affected vs neurotypical parents*** |
| --- | --- | --- | --- | --- |
| **Family Income** (≤ £19,999) | 38 (40%) | 24 (12%) | 82 (31.4%) | **χ^2^ = 28.01, df = 1, p < 1.20 x 10^-7^** |
| **Maternal Education** (university/post-graduate degree) | 39 (41.1%) | 77 (39.1%) | 87 (33.0%) | χ ^2^ = 0.04, df = 1, p = 8.46 x 10^-1^ |
| ***Family Functioning: Family Climate and Caregiver-CYP relationship*** | | | | |
|  | **Families with parent(s) affected by ND-CNV** | **Families with neurotypical parents** | **Families where parent ND-CNV is unknown** | **Statistic**  ***affected vs neurotypical parents*** |
| Cohesion  Mean **±** SD | 6.92 ± 0.83 | 7.14 ± 0.98 | 7.07 ± 0.90 | **t = 2.47, df = 315.16, p < 1.41 x 10^-2^** |
| Conflict  Mean **±** SD | 4.89 ± 1.19 | 4.45 ± 1.16 | 4.71 ± 1.16 | **t = -3.69, df = 265.64, p < 2.71 x 10^-4^** |
| Warmth  Mean **±** SD | 5.51 ± 0.59 | 5.52 ± 0.59 | 5.53 ± 0.51 | t = 0.09, df = 276.07, p < 9.25 x 10^-1^ |
| Hostility  Mean **±** SD | 1.62 ± 0.53 | 1.63 ± 0.53 | 1.62 ± 0.56 | t = 0.09, df = 272.52, p < 9.28 x 10^-1^ |

a. *63 families with an affected mother, 30 families with an affected father, 2 families where both parents are affected*
