## Supplementary Table 5 for "Family functioning and psychiatric outcomes in children and young people with intellectual and developmental disabilities caused by rare genetic mutations"

**Supplementary Table 5. Sensitivity analyses for family functioning models controlling for ND-CNV status of parents.** Table presents the findings of regression models for family functioning on psychiatric and cognitive outcomes when controlling for whether CYP were part of a family with a parent with an ND-CNV. The main effects and conclusions were unchanged. Significant associations are highlighted in bold text.

| **Model** | ***Cohesion*** | ***Conflict*** | ***Warmth*** | ***Hostility*** |
| --- | --- | --- | --- | --- |
| **ADHD** | **β= -0.11, CI: -0.21--0.02, p= 2.13 × 10^-2^** | **β= 0.23, CI: 0.13-0.32, p= 4.40 × 10^-6^** | β= -0.04, CI: -0.14-0.07, p= 4.88 × 10^-1^ | **β= 0.15, CI: 0.05-0.25, p= 2.82 × 10^-3^** |
| **ODD** | **β= -0.20, CI: -0.30--0.10, p= 1.57 × 10^-4^** | **β= 0.37, CI: 0.27-0.46, p= 7.23 × 10^-13^** | **β= -0.14, CI: -0.25--0.03, p= 9.80 × 10^-3^** | **β= 0.33, CI: 0.23-0.44, p= 4.41 × 10^-10^** |
| **Mood** | **β= -0.13, CI: -0.24--0.03, p= 1.59 × 10^-2^** | **β= 0.26, CI: 0.16-0.37, p= 1.80 × 10^-6^** | β= -0.10, CI: -0.21-0.02, p= 9.53 × 10^-2^ | **β= 0.24, CI: 0.13-0.35, p= 2.16 × 10^-5^** |
| **Anxiety** | β= -0.01, CI: -0.11-0.10, p= 9.17 × 10^-1^ | **β= 0.13, CI: 0.02-0.24, p= 2.09 × 10^-2^** | β= 0.03, CI: -0.09-0.14, p= 6.49 × 10^-1^ | β= 0.01, CI: -0.10-0.13, p= 8.06 × 10^-1^ |
| **OCD** | β= 0.06, CI: -0.06-0.18, p= 3.10 × 10^-1^ | β= 0.05, CI: -0.07-0.17, p= 4.13 × 10^-1^ | β= 0.06, CI: -0.06-0.19, p= 3.24 × 10^-1^ | β= -0.001, CI: -0.13-0.12, p= 9.82 × 10^-1^ |
| **FSIQ** | β= 0.06, CI: -1.59-1.70, p= 9.45 × 10^-1^ | β= 0.67, CI: -0.96-2.30, p= 4.19 × 10^-1^ | β= -1.02, CI: -2.69-0.65, p= 2.31 × 10^-1^ | β= 0.17, CI: -1.51-1.85, p= 8.41 × 10^-1^ |
| **PIQ** | β= 0.06, CI: -1.71-1.83, p= 9.47 × 10^-1^ | β= 1.06, CI: -0.74-2.86, p= 2.46 × 10^-1^ | β= -0.74, CI: -2.55-1.07, p= 4.21 × 10^-1^ | β= -0.45, CI: -2.29-1.39, p= 6.29 × 10^-1^ |
| **VIQ** | β= 0.14, CI: -1.57-1.84, p= 8.76 × 10^-1^ | β= 0.24, CI: -1.46-1.93, p= 7.83 × 10^-1^ | β= -1.04, CI: -2.77-0.69, p= 2.39 × 10⁻⁰¹ | β= 0.68, CI: -1.06-2.42, p= 4.43 × 10^-1^ |
