## Supplementary Table 6 for "Family functioning and psychiatric outcomes in children and young people with intellectual and developmental disabilities caused by rare genetic mutations"

**Supplementary Table 6. Sensitivity analyses for caregiver-CYP relationships with mothers only.** Table presents the findings of regression models for caregiver-CYP relationships on psychiatric and cognitive outcomes in a sample of families using mothers’ reports only. The main effects and conclusions were unchanged. Significant associations are highlighted in bold text.

| **Model** | ***Warmth*** | ***Hostility*** |
| --- | --- | --- |
| **ADHD** | β = -0.07, CI: -0.14-0.003, p = 6.06 × 10⁻² | **β = 0.17, CI: 0.10-0.24, p = 2.44 × 10⁻⁶** |
| **ODD** | **β = -0.16, CI: -0.24--0.08, p = 1.42 × 10^-4^** | **β = 0.32, CI: 0.24-0.39, p = 4.28 × 10⁻¹⁵** |
| **Mood** | **β = -0.08, CI: -0.17--0.003, p = 4.21 × 10⁻²** | **β = 0.20, CI: 0.12-0.27, p = 7.05 × 10⁻⁷** |
| **Anxiety** | β = 0.06, CI: -0.02-0.15, p = 1.51 × 10⁻¹ | β = -0.03, CI: -0.11-0.06, p = 5.27 × 10⁻¹ |
| **OCD** | β = 0.06, CI: -0.03-0.15, p = 1.91 × 10⁻¹ | β = -0.03, CI: -0.12-0.06, p = 5.19 × 10⁻¹ |
| **FSIQ** | β = -1.01, CI: -2.31-0.30, p = 1.30 × 10⁻¹ | β = 0.39, CI: -0.87-1.64, p = 5.46 × 10⁻¹ |
| **PIQ** | β = -0.32, CI: -1.70-1.06, p = 6.50 × 10⁻¹ | β = -0.04, CI: -1.37-1.29, p = 9.53 × 10⁻¹ |
| **VIQ** | β = -1.20, CI: -2.56-0.16, p = 8.32 × 10⁻² | β = 1.17, CI: -0.14-2.48, p = 7.98 × 10⁻² |
