## Supplementary Table 7 for "Family functioning and psychiatric outcomes in children and young people with intellectual and developmental disabilities caused by rare genetic mutations"

**Supplementary Table 7. Associations between family functioning, family income and maternal education.**

Table presents the findings of regression models for family income and maternal education on family functioning measures. Significant associations are highlighted in bold text.

| **Model** | **Family income** | **Maternal education** |
| --- | --- | --- |
| **Cohesion** | **β = 0.16, CI = 0.08 - 0.23, p = 5.48 × 10⁻⁵** | β = 0.18, CI = -0.14 - 0.27, p = 7.18 × 10⁻¹ |
| **Conflict** | **β = -0.23, CI = -0.31 - -0.15, p = 6.00 × 10⁻⁹** | **β = -0.42, CI = -0.75 - -0.10, p = 1.10 × 10⁻^2^** |
| **Warmth** | β = -0.02, CI = -0.09 - 0.06, p = 6.59 × 10⁻¹ | β = -0.21, CI = -0.53 - 0.11, p = 1.94 × 10⁻^1^ |
| **Hostility** | β = 0.01, CI = -0.07 - 0.09, p = 8.43 × 10⁻¹ | β = 0.04, CI = -0.29 - 0.37, p = 8.06 × 10⁻¹ |
