## Supplementary Table 8 for "Family functioning and psychiatric outcomes in children and young people with intellectual and developmental disabilities caused by rare genetic mutations"

**Supplementary Table 8. Regression models for family functioning * ND-CNV interactions and psychiatric outcomes.**

Interaction models of family functioning measures and ND-CNV status while controlling age, sex assigned at birth, family income and maternal education (Aim 5). Models were conducted with normalized z-scores for both family functioning measures and symptoms counts for psychiatric outcomes (ADHD, ODD, Mood). Associations that remained significant following correction are highlighted in bold text.

| **Family Functioning** | **Effect** | **ADHD** | **ODD** | **Mood** |
| --- | --- | --- | --- | --- |
| Cohesion | Cohesion | β = -0.022, CI: -0.134-0.09, p = 6.99 × 10⁻⁰¹ | β = -0.064, CI: -0.189-0.062, p = 3.20 × 10⁻⁰¹ | β = -0.099, CI: -0.224-0.026, p = 1.19 × 10⁻⁰¹ |
|  | Age (Years) | β = -0.031, CI: -0.05--0.012, p = 1.36 × 10⁻⁰³ | β = -0.027, CI: -0.048--0.006, p = 1.22 × 10⁻⁰² | β = 0.045, CI: 0.024-0.066, p = 3.40 × 10⁻⁰⁵ |
|  | Sex assigned at birth (females) | β = -0.229, CI: -0.346--0.111, p = 1.23 × 10⁻⁰⁴*** | β = -0.022, CI: -0.152-0.109, p = 7.45 × 10⁻⁰¹ | β = 0.024, CI: -0.105-0.154, p = 7.13 × 10⁻⁰¹ |
|  | Family income | β = -0.121, CI: -0.182--0.06, p = 1.08 × 10⁻⁰⁴ | β = -0.083, CI: -0.152--0.013, p = 2.00 × 10⁻⁰² | β = -0.101, CI: -0.173--0.029, p = 5.94 × 10⁻⁰³ |
|  | Maternal education | β = 0, CI: -0.071-0.07, p = 9.94 × 10⁻⁰¹ | β = -0.025, CI: -0.105-0.056, p = 5.48 × 10⁻⁰¹ | β = -0.038, CI: -0.121-0.046, p = 3.76 × 10⁻⁰¹ |
|  | ND-CNV | β = 1.132, CI: 1.003-1.261, p = 1.72 × 10⁻⁵³ | β = 0.743, CI: 0.601-0.886, p = 2.54 × 10⁻²² | β = 0.696, CI: 0.557-0.836, p = 7.54 × 10⁻²¹ |
|  | Cohesion*ND-CNV | β = -0.1, CI = -0.23 - 0.03, p = 1.23 × 10⁻⁰¹ | **β = -0.14, CI = -0.28 - 0, p = 4.65 × 10⁻⁰²** | β = -0.04, CI = -0.18 - 0.1, p = 5.61 × 10⁻⁰¹ |
| Conflict | Conflict | β = 0.08, CI: -0.028-0.189, p = 1.47 × 10⁻⁰¹ | β = 0.179, CI: 0.06-0.297, p = 3.19 × 10⁻⁰³ | β = 0.125, CI: 0.004-0.246, p = 4.30 × 10⁻⁰² |
|  | Age (Years) | β = -0.028, CI: -0.046--0.009, p = 3.80 × 10⁻⁰³ | β = -0.021, CI: -0.041-0, p = 4.82 × 10⁻⁰² | β = 0.049, CI: 0.028-0.07, p = 4.63 × 10⁻⁰⁶ |
|  | Sex assigned at birth (females) | β = -0.225, CI: -0.341--0.108, p = 1.44 × 10⁻⁰⁴ | β = -0.018, CI: -0.144-0.109, p = 7.85 × 10⁻⁰¹ | β = 0.03, CI: -0.098-0.158, p = 6.45 × 10⁻⁰¹ |
|  | Family income | β = -0.104, CI: -0.164--0.043, p = 7.77 × 10⁻⁰⁴ | β = -0.045, CI: -0.111-0.021, p = 1.82 × 10⁻⁰¹ | β = -0.081, CI: -0.152--0.01, p = 2.46 × 10⁻⁰²* |
|  | Maternal education | β = 0.009, CI: -0.06-0.079, p = 7.91 × 10⁻⁰¹ | β = -0.008, CI: -0.084-0.068, p = 8.40 × 10⁻⁰¹ | β = -0.026, CI: -0.108-0.055, p = 5.25 × 10⁻⁰¹ |
|  | ND-CNV | β = 1.135, CI: 1.006-1.264, p = 4.22 × 10⁻⁵⁴ | β = 0.748, CI: 0.607-0.888, p = 2.32 × 10⁻²³*** | β = 0.707, CI: 0.568-0.846, p = 1.50 × 10⁻²¹*** |
|  | Conflict*ND-CNV | β = 0.1, CI = -0.03 - 0.22, p = 1.30 × 10⁻⁰¹ | **β = 0.18, CI = 0.04 - 0.31, p = 1.05 × 10⁻⁰²** | β = 0.1, CI = -0.03 - 0.23, p = 1.36 × 10⁻⁰¹ |
| Warmth | Warmth | β = -0.068, CI: -0.177-0.041, p = 2.19 × 10⁻⁰¹ | β = -0.075, CI: -0.198-0.047, p = 2.27 × 10⁻⁰¹ | β = -0.052, CI: -0.173-0.07, p = 4.01 × 10⁻⁰¹ |
|  | Age (Years) | β = -0.034, CI: -0.053--0.015, p = 5.25 × 10⁻⁰⁴ | β = -0.033, CI: -0.055--0.011, p = 2.92 × 10⁻⁰³ | β = 0.041, CI: 0.02-0.063, p = 2.14 × 10⁻⁰⁴ |
|  | Sex assigned at birth (females) | β = -0.231, CI: -0.349--0.113, p = 1.11 × 10⁻⁰⁴ | β = -0.028, CI: -0.158-0.103, p = 6.79 × 10⁻⁰¹ | β = 0.023, CI: -0.107-0.153, p = 7.29 × 10⁻⁰¹ |
|  | Family income | β = -0.136, CI: -0.196--0.076, p = 1.14 × 10⁻⁰⁵*** | β = -0.109, CI: -0.178--0.039, p = 2.33 × 10⁻⁰³** | β = -0.122, CI: -0.193--0.051, p = 8.29 × 10⁻⁰⁴*** |
|  | Maternal education | β = -0.007, CI: -0.078-0.065, p = 8.52 × 10⁻⁰¹ | β = -0.035, CI: -0.118-0.048, p = 4.08 × 10⁻⁰¹ | β = -0.045, CI: -0.13-0.039, p = 2.93 × 10⁻⁰¹ |
|  | ND-CNV | β = 1.135, CI: 1.006-1.265, p = 1.33 × 10⁻⁵³ | β = 0.748, CI: 0.606-0.89, p = 9.38 × 10⁻²³ | β = 0.704, CI: 0.565-0.843, p = 2.31 × 10⁻²¹* |
|  | Warmth*ND-CNV | β = 0, CI = -0.13 - 0.12, p = 9.87 × 10⁻⁰¹ | β = -0.07, CI = -0.21 - 0.07, p = 3.48 × 10⁻⁰¹ | β = -0.04, CI = -0.18 - 0.1, p = 5.56 × 10⁻⁰¹ |
| Hostility | Hostility | β = 0.079, CI: -0.037-0.194, p = 1.81 × 10⁻⁰¹ | β = 0.215, CI: 0.089-0.341, p = 8.08 × 10⁻⁰⁴ | β = 0.079, CI: -0.049-0.207, p = 2.22 × 10⁻⁰¹ |
|  | Age (Years) | β = -0.033, CI: -0.052--0.015, p = 4.82 × 10⁻⁰⁴ | β = -0.032, CI: -0.053--0.012, p = 2.02 × 10⁻⁰³ | β = 0.042, CI: 0.021-0.063, p = 1.03 × 10⁻⁰⁴ |
|  | Sex assigned at birth (females) | β = -0.228, CI: -0.344--0.112, p = 1.08 × 10⁻⁰⁴ | β = -0.017, CI: -0.142-0.108, p = 7.86 × 10⁻⁰¹ | β = 0.033, CI: -0.094-0.16, p = 6.11 × 10⁻⁰¹ |
|  | Family income | β = -0.136, CI: -0.196--0.077, p = 8.37 × 10⁻⁰⁶ | β = -0.107, CI: -0.173--0.04, p = 1.76 × 10⁻⁰³ | β = -0.122, CI: -0.192--0.052, p = 6.95 × 10⁻⁰⁴ |
|  | Maternal education | β = 0.001, CI: -0.069-0.072, p = 9.70 × 10⁻⁰¹ | β = -0.022, CI: -0.101-0.057, p = 5.77 × 10⁻⁰¹ | β = -0.037, CI: -0.12-0.046, p = 3.81 × 10⁻⁰¹ |
|  | ND-CNV | β = 1.142, CI: 1.016-1.269, p = 1.46 × 10⁻⁵⁵ | β = 0.77, CI: 0.634-0.905, p = 1.00 × 10⁻²⁵ | β = 0.717, CI: 0.582-0.854, p = 8.38 × 10⁻²³ |
|  | Hostility*ND-CNV | β = 0.1, CI = -0.03 - 0.23, p = 1.43 × 10⁻⁰¹ | β = 0.12, CI = -0.02 - 0.26, p = 1.03 × 10⁻⁰¹ | **β = 0.15, CI = 0 - 0.29, p = 4.55 × 10⁻⁰²** |
